# Using chest X-ray to screen for Tuberculosis on arrival to prison: A service evaluation

**DOI:** 10.1101/2025.04.11.25325539

**Authors:** S E Perrett, M Backx, E Lewandowski, R Lightburn, S Roberts, T Rooke, G Ahern, B J Gray

## Abstract

**Objectives:** Chest X-ray (CXR) is recommended by the World Health Organization as a TB screening tool on admission to prison. We piloted the offer of CXR on admission to prison to understand if it was acceptable to residents, feasible to deliver within 48 hours of admission, and to inform TB epidemiology.

**Study Design:** Cross-sectional

**Method:** Between 1 September 2023 and 31 March 2024, CXRs were offered to new prison admissions. We measured the numbers accepting, numbers completed and the results. For each person accepting the CXR we undertook an assessment of clinical and social TB risks. We measured the time taken to deliver the CXR and receive results.

**Results:** CXR was acceptable to those in prison with 61% (n=310) of new admissions accepting the offer. Of those accepting the offer, 226 (73%) went on to receive a CXR, equating to 44% of all new arrivals within the pilot period. A quarter of those accepting the CXR offer did not attend their first appointment and needed further appointment offers. We observed that as the number of rearranged appointments increased the number of men attending decreased. The total median number of days from arrival at the prison to completion of CXR was 17 [IQR 13-20 days]. We did not identify any respiratory TB, however nine (4%) CXRs were abnormal..

**Conclusion:** CXR screening was acceptable to prison residents but we could not achieve delivery within 48 hours of arrival to prison. We identified other respiratory abnormalities suggesting CXR screening could be used as a wider respiratory health screen of which TB would be included.

**What this study adds:**

- CXR screening for TB on admission to prison is recommended despite a low certainty of evidence for this approach. Our findings contribute to the evidence and demonstrate the complexity of implementing this in practice
- The offer of CXR on admission to prison was acceptable to new arrivals although delivery within 48 hours of arrival to prison could not be achieved within the existing service model
- Whilst we did not identify any TB, we identified other respiratory abnormalities suggesting CXR screening on arrival could be used as a wider respiratory health screen.

**Implications for policy and practice:**

- To achieve CXR within 48 hours of admission to prison would require a bespoke service model in addition to existing prison radiography provision
- Policies should consider CXR TB screening as part of a wider respiratory health screen, offering the opportunity to address multiple health issues at the same time
- The frequent movement of people through the criminal justice system presents a challenge to continuity of care and should be considered when designing interventions

## INTRODUCTION

The World Health Organization (WHO) End TB Strategy aims to reduce TB incidence by 80% by 2030^1^. As the WHO acknowledges, achieving this will be dependent on bridging the case-detection gap amongst populations most vulnerable to TB, including those in prison. Prison populations are known to present late, suffer worse health outcomes and, due to closely confined living conditions, are at high risk of respiratory infection transmission^2,3^. TB screening aims to detect disease early, improve opportunity for timely treatment and reduce the risk of onward transmission^4^.

To improve respiratory TB screening amongst prison populations, WHO advise chest X-ray (CXR) as a single screening algorithm. UK clinical guidance supports this, recommending that prisons with CXR facilities offer CXR within 48 hours of arrival^5^. Current screening in most UK prisons is via symptom screening or patient presentation with variable implementation. Very few UK prisons have access to CXR on site.

Despite recommending CXR screening for prison populations, the WHO recognise there is a low certainty of evidence for this approach and that monitoring, and evaluation should be part of any screening programme^4^. We worked with a UK prison that has permanent on-site CXR facilities to pilot CXR as a single TB screening algorithm for new arrivals. We sought to answer three questions: (i) was the offer of CXR screening acceptable to those in prison? (ii) Was it feasible to implement CXR as a single screening algorithm for TB in the prison environment? (iii) What does CXR screening for new arrivals tell us about the epidemiology of TB and TB risk at the prison?

## METHODS

### Setting

The service pilot took place at a secure training and resettlement prison in Wales, UK. The prison holds approximately 2,000 men and receives around 40 new arrivals each week. The prison is managed by His Majesty’s Prison and Probation Service (HMPPS), an executive agency of the UK Ministry of Justice. A dedicated multidisciplinary health and wellbeing service is provided on-site by the National Health Service (NHS) and includes static X-ray facilities.

### Screening Algorithm

A TB CXR screening algorithm, in line with the model proposed by the WHO^1^, was developed and offered CXR to new arrivals between 1 September 2023 and 31 March 2024. Healthcare staff were trained to make the offer as part of the new arrival health screen. Information leaflets were given to each new arrival explaining why the CXR was being offered, what Tuberculosis is, what would happen if something was found on the CXR, risks of having the CXR and assurance that the CXR offer was optional. Anyone accepting the CXR offer was referred by the prison medical practitioner to the prison radiographer. CXR sessions were held at the prison during three half day sessions per week. CXRs were read by the radiography team at the local hospital as per normal health service processes at the prison. Results were returned to the prison within 2-3 weeks. Clinical pathways were agreed with the specialist respiratory team at the local hospital for referral of any abnormal CXRs.

### Measures

We measured acceptability as the proportion of men accepting the screening offer on arrival at the prison.

To measure feasibility, we considered the proportion screened by CXR against the proportion offered. To further understand feasibility of service delivery we measured the time between arrival at the prison and screening offer, the time between acceptance and completion of the CXR, the number of CXR appointments offered before attendance and finally the time for CXR results to be reported back to the prison. We measured these by median days and interquartile range.

To understand the epidemiology of TB we measured uptake and abnormal results as a percentage. To understand TB risk, a risk assessment questionnaire was undertaken for each person accepting the CXR offer. This included age group, ethnicity, country of origin, history of homelessness, history of smoking, history drugs and alcohol misuse, and any known contact with a TB case. To understand clinical risk, we asked about history of cough, weight loss, night sweats, fever, loss of appetite and previous TB illness.

### Data Analysis

Descriptive analysis was undertaken initially to provide counts and proportions. Chi-squared analyses was used to examine the association between the two groups: individuals who had a chest X-ray (CXR) and those who did not. For the service delivery aspects, data is presented as median days and interquartile ranges

## RESULTS

### Acceptability and clinical findings

Between 1 September 2023 and 31 March 2024, 508 men arrived at the prison. Of these, 310 (61%) accepted the offer of CXR screening for TB. The cohort accepting the CXR offer were young, 69% were aged 39 years or younger, 87% were White and only 6.5% born outside of the UK. The majority had previous experience of being in prison (71%) (Table 1). Acceptance of CXR screening was lower in those arriving from another prison (59%) compared to those arriving directly from the community (69%) (Table 1).

**Table 1.** Demographic and social risk factors of those accepting the CXR offer.

| Demographic or Social Risk Factor | Remand<br>(n=77) | Transfer<br>(n=233) | All<br>(n=310) |
| --- | --- | --- | --- |
| **18-29 years** | 20 (20.6%) | 76 (32.6%) | 96 (31.0%) |
| **30-39 years** | 30 (39.0%) | 87 (37.3%) | 117 (37.7%) |
| **40-49 years** | 19 (24.7%) | 53 (22.7%) | 72 (23.2%) |
| **50 years or over** | 8 (10.4%) | 16 (6.9%) | 24 (7.7%) |
| **White** | 69 (89.6%) | 200 (85.8%) | 269 (86.8%) |
| **Global majority** | 8 (10.4%) | 31 (13.2%) | 39 (12.6%) |
| **UK Born** | 69 (89.6%) | 220 (94.4%) | 289 (93.2%) |
| **Non-UK Born** | 8 (10.4%) | 12 (5.2%) | 20 (6.5%) |
| **Previous prison stay** | 37 (48.1%) | 182 (78.1%)* | 219 (70.6%) |
| **Smoker before prison/currently vaping** | 59 (76.6%) | 190 (81.5%) | 249 (80.3%) |
| **Alcohol misuse** | 20 (26.0%) | 56 (24.0%) | 76 (24.5%) |
| **Smoked drugs - Never** | 57 (74.0%) | 125 (53.6%)* | 182 (58.7%) |
| **Smoked drugs – Current (user before prison)** | 7 (9.1%) | 22 (9.4%)* | 29 (9.4%) |
| **Smoked drugs – Recent (in past 5 years)** | 11 (14.3%) | 63 (27.0%)* | 74 (23.9%) |
| **Smoked drugs – Past (more than 5 years ago)** | 2 (2.6%) | 17 (7.3%)* | 19 (6.1%) |
| **Injected drugs - Never** | 68 (88.3%) | 183 (78.5%) | 251 (81.0%) |
| **Injected drugs – Current (user before prison)** | 2 (2.6%) | 9 (3.9%) | 11 (3.5%) |
| **Injected drugs – Recent (in past 5 years)** | 5 (6.5%) | 21 (9.0%) | 26 (8.4%) |
| **Injected drugs – Past (more than 5 years ago)** | 1 (1.3%) | 15 (6.4%) | 16 (5.2%) |
| **Time homeless - None** | 41 (53.2%) | 137 (58.8%)* | 178 (57.4%) |
| **Time homeless - Less than a year** | 24 (31.2%) | 38 (16.3%)* | 62 (20.0%) |
| **Time homeless - 1 year** | 7 (9.1%) | 24 (10.3%)* | 31 (10.0%) |
| **Time homeless - 2-3 years** | 2 (2.6%) | 14 (6.0%)* | 16 (5.2%) |
| **Time homeless - More than 3 years** | 2 (2.6%) | 19 (8.2%)* | 21 (6.8%) |
| **Close contact with TB case** | 5 (6.5%) | 9 (3.8%) | 14 (4.5%) |

Of 310 men accepting the screening offer, 226 (73%) went on to receive a CXR, equating to 44% of all new arrivals within the pilot period. We did not identify any cases of respiratory TB however, nine (4%) CXRs were marked as abnormal, requiring follow up. Of these, six (67%) were successfully followed up and no further action was required, two (22%) were released before follow up and one declined follow up.

Of the abnormal CXR results, four were reported as “? Chronic Obstructive Pulmonary Disease (COPD) changes” or “Chronic post inflammatory change” and were followed up by the prison physician. All four were given health promotion advice around smoking but no further follow up was required. A further four CXR results advised a CT scan of which two patients (“?inflammation of left lung” and “?pulmonary contusion”) received a CT but no further action was required, the remaining two (“CT abdo/pelvis advised” and “Recommend CT chest”) were released before a CT could be arranged and were advised to follow up in the community. One CXR result advised follow up due to “cardiac indications enlarged” however this patient declined any further review.

### Feasibility

The median number of days between arrival at the prison and referral for CXR was 5 [IQR 3-7 days]. The median number of days from referral to completion of the CXR was 12 [IQR 8-16 days]. The total median number of days from arrival at the prison to completion of CXR was 17 [IQR 13-20 days]. Following CXR, it took a median of 7 days for the CXR to be reported back to the prison [IQR 3-8 days].

A quarter of those accepting the CXR offer did not attend their first appointment and needed further appointment offers (Figure 1). As the number of rearranged appointments increased the number of men attending decreased, in a linear trend (Figure 1).

**Figure 1.**
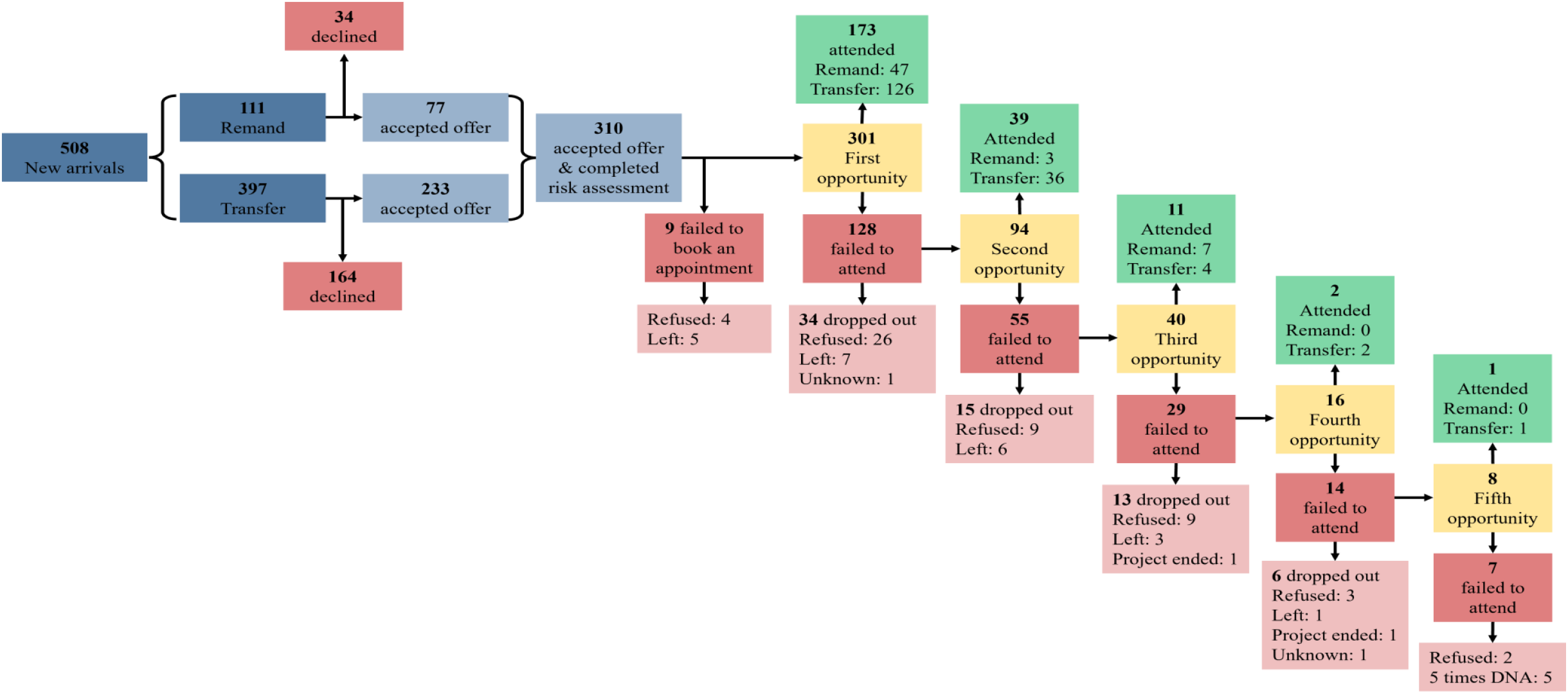
Flow of individuals through appointment process for CXR.

Of the nine abnormal CXRs needing clinical follow up, four were released before being able to be reviewed.

### Social and clinical risk factors for TB

We identified an abundance of social risk factors amongst those accepting CXR screening. Just over a third had a history of having smoked illicit drugs (39%) and 17% had injected illicit drugs. Just under half (42%) had experience of homelessness. In terms of a risk factor for broader respiratory health, 80% had previously smoked or vaped (Table 1). The more social risk factors, the higher the tendency of not completing a CXR. Of those with no identified social risk factors, 78% completed their CXR compared to 61% of those with five or more social risk factors (Figure 2).

**Figure 2.**
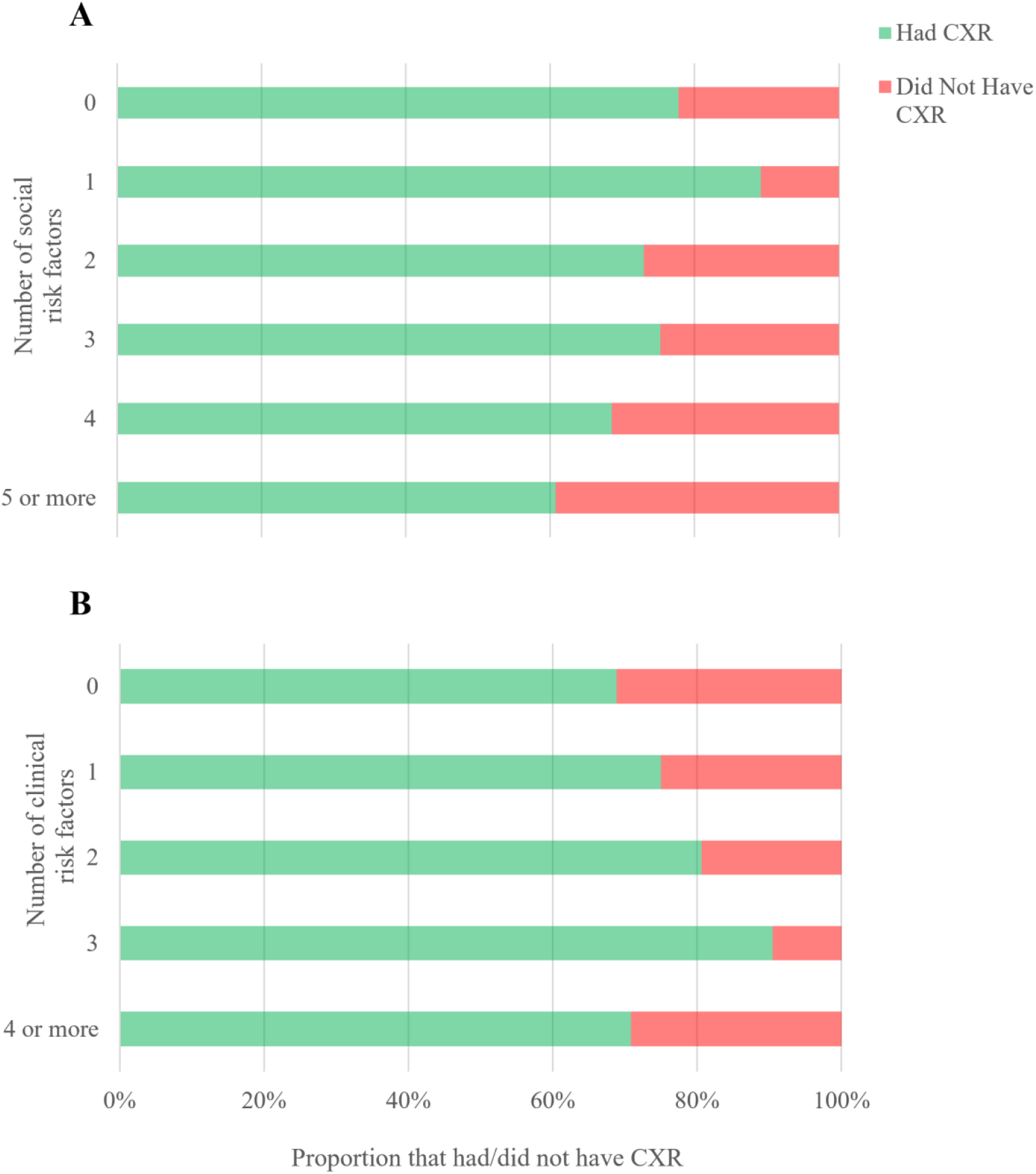
The proportion of those who had or did not have a chest X-ray in relation to (A) the number of social and (B) the number of clinical risk factors The social risk factors were (i) history of smoking drugs, (ii) history of injecting drugs, (iii) history of homelessness, (iv) smoker prior to prison entry and/or currently vaping, (v) previous prison stay, (vi) history of alcohol abuse and (vi) global majority ethnicity and/or country of birth outside UK.

The most common clinical risk factor experienced by those accepting the CXR was night sweats (26%) followed by loss of appetite (21%) and persistent cough (19%) (Table 2). The tendency of completing a CXR increased for those who had 0-3 clinical risk factors. Seventy percent of those with no clinical risk factors completed a CXR compared to 90% of those with three clinical risk factors (Figure 2).

**Table 2.**
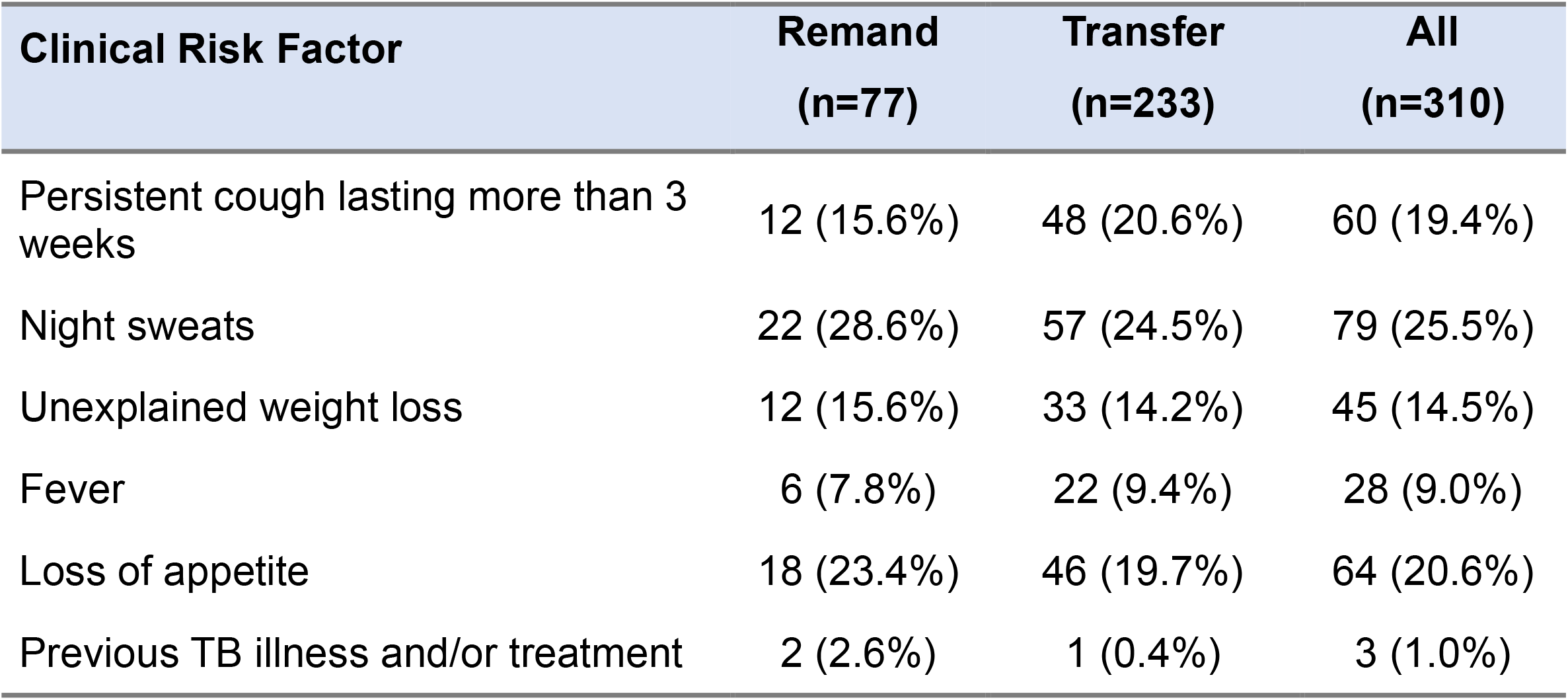
Clinical risk factors amongst new arrivals remanded and those transferred from another prison.

We found a higher number of abnormal CXRs (n=8) in those with three or fewer social risk factors although one abnormal CXR had five or more social risk factors. There was no clear relationship between the number of clinical risk factors and abnormal CXRs (Table 3).

**Table 3.** Social and clinical risk factors amongst those screened and those with abnormal CXR results.

| Number of social risk factors | Number of screened | Number with abnormal CXR | Percentage abnormal CXR |
| --- | --- | --- | --- |
| 0 | 7 | 0 | 0.0 |
| 1 | 33 | 0 | 0.0 |
| 2 | 51 | 3 | 5.6 |
| 3 | 59 | 5 | 7.8 |
| 4 | 37 | 0 | 0.0 |
| 5 or more | 30 | 1 | 3.2 |

| Number of clinical risk factors | Number screened | Number with abnormal CXR | Percentage abnormal CXR |
| --- | --- | --- | --- |
| 0 | 126 | 3 | 2.4 |
| 1 | 39 | 2 | 5.1 |
| 2 | 25 | 1 | 4.0 |
| 3 | 19 | 1 | 5.3 |
| 4 or more | 17 | 2 | 11.8 |

## DISCUSSION

Just under two-thirds of all new arrivals to the prison accepted the offer of CXR screening for Tuberculosis demonstrating the intervention was acceptable to most men. Despite initially high levels of engagement, a quarter of men did not attend their first appointment and after three appointment offers men were very unlikely to attend. Less than half of new admissions went on to receive a CXR. Further enquiry is needed to understand if non-attendance is due to logistics such as clashes with other commitments such as education, work or visits, or staff escort shortages, or whether it reflects a real reduction in levels of engagement.

Our measures of the time taken to deliver the service highlight operational challenges to delivering CXRs in a prison environment. We were unable to achieve screening within 48 hours of admission, as per advice from NICE^5^, for anyone who took part in the pilot. It commonly took over two weeks (median of 17 days) from arrival to complete a CXR. Time was needed to ensure people were given adequate information, consented, issued with an appointment and facilitated to attend. The first 48 hours of arrival at prison are administratively busy requiring multiple assessments from both health and justice sectors. Those in prison are known to suffer from poor health and many men may present with acute physical or mental health issues that take priority on arrival. We delivered the screening within the existing radiography service model. CXR sessions were held during office hours and we did not have additional resource to escort residents to and from the CXR facility. Although not measured as part of this pilot, logistical barriers, such as CXR facilities being in a different area of the prison to arrivals, security escorts to move men around the prison, and arrivals commonly happening late in the afternoon/evening will inevitably impact the timeliness of delivering this health intervention. The time required to deliver the service supports previously described logistical challenges to delivering healthcare in a gatekeeping system as found in the literature^6,7,8^. A bespoke service model is likely to be needed to achieve CXR within 48 hours of admission to prison.

It took a further week for CXR results to be returned to the prison and although not measured, it can be assumed that more administrative time is required to check these results and book men into follow up appointments. Advances in artificial intelligence algorithms to detect TB from CXRs may enable results to be delivered more swiftly in the future.^9^ This may be particularly useful for high-risk mobile populations. Two men with abnormal results were released before they could be followed up demonstrating the challenges of ensuring continuity of care to a mobile population with high churn.

We did not identify any TB disease. Wales remains within the WHO definition of a low incidence country with annual incidence at 2.7 per 100,00 population^10^. The prevalence of active TB disease amongst prison populations in Wales is not known. A previous screening pilot at a prison in Wales found latent TB positivity of 7.1% and active TB of 0.1%^11^. Based on previous estimates of number needed to screen (NNS) its possible we did not screen enough people to be able to find a single case^12^.

Despite not identifying TB disease we found a high level of TB risk factors (social and clinical) supporting the evidence that this population remains at elevated risk of TB disease. Concerningly, the higher the number of social risk factors the lower the proportion who attended CXR. Qualitative enquiry would be useful to understand factors influencing this relationship. The increase in social risk factors could indicate increased transiency and reduced likelihood to engage in health interventions in general but there may also be social barriers to attending or issues relating to stigma that need to be understood. Methods to better engage with those at increased social risk are needed to screening programmes reach those at greatest risk.

Conversely, the more clinical risk factors reported the higher the proportion who attended CXR. It could be that those who feel physically unwell are more likely to actively engage with diagnostic interventions. We saw a drop off at four clinical risk factors that needs exploring further.

We did not find a clear relationship between the number of risk factors and abnormal CXR results.

Although no TB was identified, the CXR screening identified nine possible clinical abnormalities for follow up. Although those followed up did not lead to any new diagnoses or treatment, health promotion advice was able to be given particularly for results suggestive of COPD. Although prisons in England and Wales are smoke-free^13^, prison populations are known to have high rates of smoking and second-hand smoking, increasing the risk for COPD. Eighty percent of our cohort had a history of smoking and just under 40% had smoked illicit substances heightening the risk of respiratory disease. The prevalence of COPD has been found to be higher in prison populations^14,15^ and respiratory disease remains a leading cause of death for imprisoned populations^16^. The pilot demonstrates that CXR screening for TB in this population may have wider benefits and could be integrated into a broader respiratory health screen.

### Limitations

At the time of the study, the prison system was experiencing significant pressures, especially in relation to capacity. Therefore, the number of men who were new arrivals was lower than previously anticipated. The high rates of churn also resulted in some men being released before they had the opportunity to attend an appointment for a CXR. However, these are the implications of undertaking any service evaluation within the prison environment and it is likely the pressures will be ongoing. The data presented is quantitative, the evaluation would have benefited from some qualitative insights especially to understand why some men although are initially interested in a CXR never end up attending an appointment. We were unable to undertake a full economic analysis of the pilot with the data provided to us by the prison healthcare team and from the health board, future research should prioritise a detailed economic evaluation of CXR screening for TB in prison.

## CONCLUSION

CXR screening for TB in prison populations is acceptable to prison residents and can be delivered successfully however we could not achieve delivery within 48 hours of arrival. Such timescales should be reserved for those with clinical suspicion of TB disease and where symptoms such as cough risk spread of infection. Our results confirmed this population carry many risk factors for TB disease and should remain prioritised for CXR screening. We identified other respiratory abnormalities suggesting CXR screening on arrival could be used as a wider respiratory health screen of which TB would be included.

## Data Availability

All data produced in the present work are contained in the manuscript

## ACKNOWLEDGEMENTS

Pam Lloyd, Stephen Kelly, Tabitha Kavoi, Aimée Challenger, Josie Smith, Richard Firth, Hannah Beer

## STATEMENT OF ETHICAL APPROVAL

Ethical oversight of the pilot and evaluation was provided by Public Health Wales NHS Trust Research & Development Office. The Public Health Wales NHS Trust Research & Development Office reviewed the project outline and determined this project was not defined as an NHS research project, according to the NHS definition of research, outlined in clause 3.1 of the UK Policy Framework for health and social care research. As a result of this decision, NHS research ethics approval was not required for this study. Public Health Wales received anonymous data and held and processed this under Public Health Wales information governance arrangements, in compliance with the Data Protection Act, Caldicott Principles and Public Health Wales guidance on the release of small numbers. A Data Protection Impact Assessment (DPIA) was completed.

## Notes

### Competing Interest Statement

The authors have declared no competing interest.

### Funding Statement

This study did not receive any funding

